# Growth Rate Assessed by Vascular Deformation Mapping predicts Type B Aortic Dissection in Marfan Syndrome

**DOI:** 10.1101/2024.10.10.24315133

**Authors:** Carlos Alberto Campello Jorge, Prabhvir Singh Marway, Nicasius S Tjahjadi, Heather A Knauer, Himanshu J Patel, Marion Hofmann Bowman, Kim Eagle, Nicholas S. Burris

**Affiliations:** Department of Radiology, University of Michigan, Ann Arbor, Michigan; Department of Cardiac Surgery, University of Michigan, Ann Arbor, Michigan; Division of Cardiovascular Medicine, Department of Internal Medicine, University of Michigan, Ann Arbor, Michigan

**Keywords:** type B aortic dissection, vascular deformation mapping, Marfan syndrome, growth rate, computed tomography angiography

## Abstract

**Background:** Patients with Marfan syndrome (MFS) are at a high risk of type B dissection (TBAD). Aortic growth and elongation have been suggested as risk factors for TBAD. Vascular deformation mapping (VDM) is an image analysis technique for mapping 3D aortic growth on rouine computed tomography angiography (CTA) scans. We aimed to use VDM to examine the value of aortic growth rate in the descending thoracic aorta (DescAo), among other imaging biomarkers, to identify the factors associated with risk of TBAD in MFS.

**Methods and Results:** CTA scans spanning 2004-2023 from adult MFS patients with native DescAo were analyzed by VDM. Other measurements included multi-level thoracoabdominal aortic diameters and the length of the DescAo by centerline analysis.

Among the 105 MFS patients analyzed, 63.8% were male, with median age of 40 years (range 18-73) and a median surveillance interval of 5.3 years (range 2.0-18.3). During surveillance, 12 (11.4%) patients developed TBAD. Patients with TBAD had higher radial growth rate (0.63 vs. 0.23 mm/year; *p* < 0.001) and elongation rate (2.4 vs. 0.5 mm/year; *p* < 0.001), on univariate and multivariable analysis, but pre-dissection descending aortic diameter was not significantly different. Predictors of growth rate included younger age, higher baseline maximal diameter of the DescAo, smoking history and warfarin use.

**Conclusions:** Radial growth and elongation rates of the DescAo were independent predictors of TBAD occurrence in MFS. TBAD often occurred in at non-aneurysmal diameters (<4.0 cm). These findings emphasize the role of growth over absolute diameter in risk stratification for TBAD in MFS.

## INTRODUCTION

Marfan syndrome (MFS) is an inherited connective tissue disorder that affects approximately 1 in 4,000 people, attributable to to gene variants within fibrillin-1. While cardiovascular manifestations drive morbidity and mortality in MFS, disease severity can vary widely between individuals.^1, 2^ Advances in prophylactic aortic root/ascending repair, along with increased rates of diagnosis, has led to an increased life expectancy by over 25 years.^3^ However, MFS patients continue to face new challenges due to longer life expectancies and extended natural disease progression, such as the risk of type B dissection (TBAD) and the development of secondary complications.^3–7^ This has resulted in a higher prevalence of chronic dissection of the distal aorta, necessitating more operations and increased morbidity and mortality.^3–5, 8–10^ Progressive dilation of the distal aorta elevates the risk of TBAD, even when the degree of dilation is mild.^6, 7, 11^ The underlying mechanisms contributing to this heightened risk of TBAD remain uncertain, with abnormal blood flow, stiffness mismatch between graft and native tissue in patients with prior root/ascending replacement, and arch geometry all playing a potential role.

Current risk assessment and management strategies in MFS rely heavily on 1-dimensional diameter measurements. However, these measurements are performed in a limited number of anatomic locations and are prone to significant variability,^12, 13^ making the confident determination of magnitude and extent of growth during surveillance very challenging. Imaging biomarkers such as the aortic tortuosity index, vertebral artery tortuosity, and the presence of primary non-aortic lesions have been explored for their potential to predict aortic growth rate and aortic events.^14–19^ Furthermore, studies have examined geometric features and conducted three-dimensional (3D) statistical analyses of thoracic aortic aneurysm shapes and geometry,^14, 15, 20^ in an attempt to establish clear links to disease etiology, severity, and clinical outcomes. However, owing to significant heterogeneity in aortic disease severity in MFS and the fact that these metrics have largely not been shown to predict key complications such as dissection, their impact on disease management remains a topic of investigation.^21, 22^

Vascular deformation mapping (VDM) is an emerging medical image analysis technique for accurate, 3D mapping of aortic growth between clinical electrocardiogram-gated CT angiography (CTA) scans. The accuracy and reproducibility of VDM technique has been validated using both clinical data and synthetic phantoms and has been shown to outperform expert raters in the identification of the location and magnitude of aortic growth. ^23^ Owing to its volumetric nature, VDM has the ability to clearly depict distinct patterns of thoracic aortic growth, including growth outside of the maximally dilated segment, among diverse cohorts of patients with hereditable thoracic aortic diseases and sporadic thoracic aortic aneurysms.^24, 25^ Understanding aortic growth rate in patients with MFS may better help define disease severity and predict complications such as TBAD.^7^

In this study, we employed VDM to examine growth of the descending thoracic among a large cohort of MFS with high-quality, longitudinal CTA imaging. We specifically aimed to investigate patient characteristics, anatomic features and 3D aortic growth metrics that are associated with development of TBAD in MFS.

## METHODS

### Study design, Population, and Outcomes

We conducted a single-center retrospective cohort study spanning February 2004 until October 2023. The institutional review board approved the study (HUM00133798) and waived the need for informed consent due to the study’s retrospective nature. We identified patients via existing aortic research databases, along with an electronic medical record search tool (DataDirect). We enrolled adult patients (>18 years) diagnosed with MFS by Ghent nosology criteria, who had at least 2 electrocardiogram-gated CTAs, with a minimum interval of 2 years to allow for detection of slow growth. The exclusion criteria were: (1) inadequate aortic enhancement (<250 Hounsfield Units); (2) thick CT slices (>2mm); (3) motion artifacts (e.g., pulsation or respiratory artifacts), which could compromise aortic segmentation quality; (4) prior surgical repair of the descending aorta; (5) a baseline diagnosis of TBAD; (6) residual dissection involving the descending thoracic aorta after prior repaired type A aortic dissection (TAAD). Baseline demographics and comorbidities were extracted through a comprehensive review of medical records. The data that support the findings of this study are available from the corresponding author upon reasonable request.

Our primary outcome of interest was TBAD, defined as the occurrence of a first non-iatrogenic TBAD. Additionally, we sought to identify patient factors that predicted growth rate as a secondary outcome. Imaging surveillance interval was defined as the period between first and last CTA used in VDM, whereas clinical follow-up interval was defined as the period between the first CTA to the last recorded patient encounter. Cardiovascular death was defined as death resulting from aortic disease, coronary artery disease, heart failure or arrhythmia.

### Standard imaging biomarkers

Standard anatomic metrics were collected on clinical CTA exams using centerline and multiplanar reformats (Vitrea, Vital Images Inc., Product Version 7.14, Minnetonka, MN, USA) thoracoabdominal aortic maximum diameters at multiple levels (thoracic: proximal, mid, and distal descending; abdominal: celiac, superior mesenteric artery, superior renal artery, inferior renal artery), as well as descending aortic length/tortuosity index, and aortic arch angle of both first and last available electrocardiogram-gated CTA. The landmarks for the diameter measurements are described in **Supplementary Figures 1-2**. The aortic arch angle was defined as the angle between the highest point of the aortic arch’s centerline and a centerline point of the ascending and descending aorta at the level of the main pulmonary artery. Descending aortic length was measured as the centerline length from immediately distal to the left subclavian artery to the proximal aspect of the celiac trunk (**Supplementary Figure 3**). The tortuosity index of the descending thoracic was quantified as the descending aortic centerline length divided by the shortest linear distance between the centerline end points.^26^ In addition to the absolute measurements, we assessed changes in the length, tortuosity, and arch angle between the two CTA images, and further normalized by the time interval (per year). Change in length by centerline is further termed “elongation”. The presence or absence of thoracoabdominal aortic calcification was assessed on baseline clinical CTA scans.

### Radial growth rate (VDM)

Radial aortic growth (perpendicular to the wall) was assessed by VDM. VDM is an emerging image analysis technique that provides precise evaluation of thoracic aortic growth using deformable image registration (Elastix 5.0.1; Utrecht University) to measure 3D deformations of the aortic wall surface occurring between two CTA scans acquired from the same individual at different time points. VDM analysis includes the following steps: (1) semi-automated segmentation of the aorta on both CTAs using a combination of an in-house neural network (U-net)^27^; (2) implicit alignment of the aortic centerline using a highly regularized multi-image, multi-metric deformable registration that applies a penalty term to enforce rigid movement of voxels within the aortic segmentation but allows deformation of periaortic structures; (3) multi-resolution, multi-metric deformable image registration using mutual information with a bending energy penalty; (4) generation of a polygonal mesh of the aortic surface from the baseline CTA geometry; (5) translation of baseline aortic mesh vertices using the deformation field calculated in step 3; (6) quantification of aortic growth as the deformation magnitude (in millimeters) of vertex-wise displacement perpendicular to the 3D aortic surface (i.e. radial direction) with color visualization in Paraview 5.8.0 (Kitware). VDM growth values are normalized by time interval to yield growth rate (mm/year). Growth rate data was then extracted from regions of the aortic mesh using Paraview, with a simplified schematic of the VDM analysis shown in **Figure 1**.

**Figure 1:**
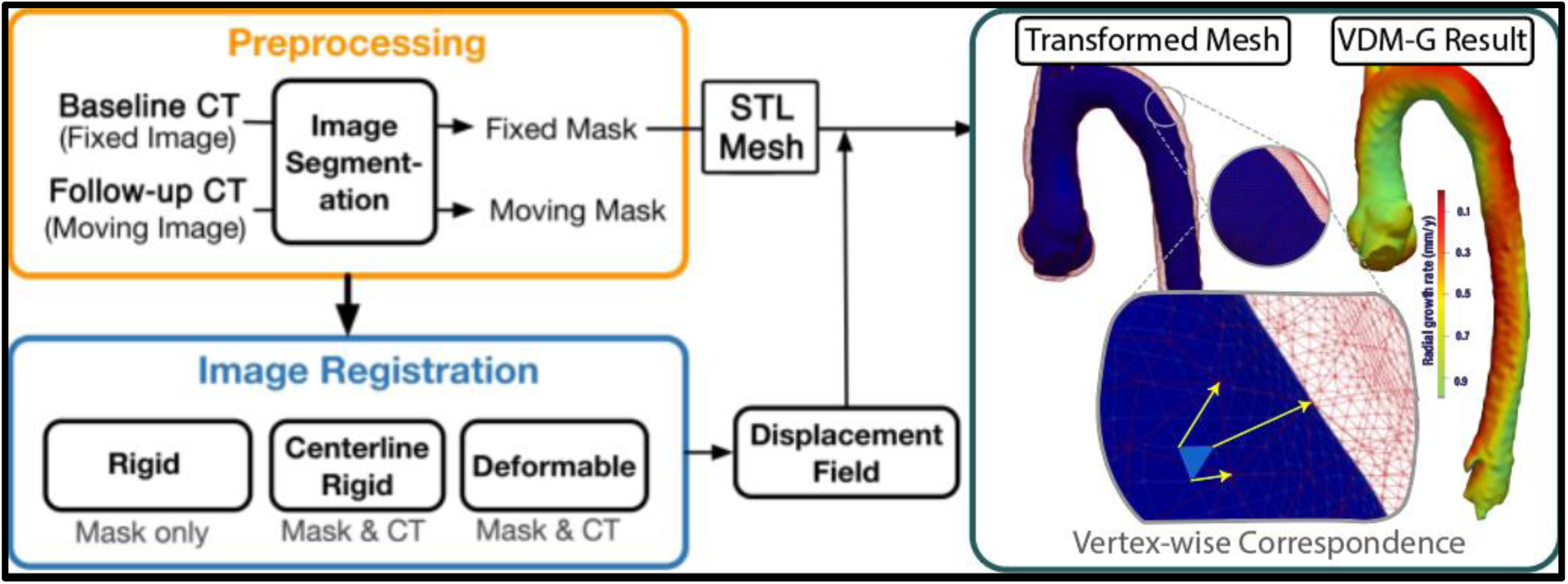
Simplified workflow involved in the vascular deformation mapping (VDM) growth mapping analysis. ECG gated aortic CTA images are retrieved for baseline and follow-up examinations, and undergo aortic segmentation (orange box), followed by rigid and deformable registration (blue box). The displacement field calculated from registration steps is used to translate the mesh vertices of the baseline model (blue surface) to the aortic geometry at follow-up (red mesh), and the deformation in the normal direction relative to the aortic surface is plotted on the 3D aortic model using a colorized scale. VDM-G = vascular deformation growth mapping. STL = stereolithography.

### Statistical Analysis

Continuous variables were expressed as means ± standard deviations or as medians with interquartile ranges (25^th^-75^th^), whereas categorical variables were expressed as percentages. Distribution of continuous variables was tested for skewness using the Shapiro-Wilk test and graphically displayed. We used Student *t* test and the Mann-Whitney *U* test to compare continuous variables, as appropriate. For comparisons of multiple groups, we performed one-way ANOVA, and we used the Kruskal-Wallis test for non-Gaussian distributions. The Fisher’s exact test was used to examine differences in frequency for categorical variables and Pearson correlation coefficients were used to assess correlation between continuous variables. Descriptive results were presented grouped by the presence of the primary outcome, and we also presented results based on sex, the presence of thoracoabdominal calcification, and prior aortic repair status. Multivariable logistic and linear regression analyses were performed to determine the independent contributions of baseline clinical characteristics and imaging biomarkers to the occurrence of TBAD at follow-up and growth rate (VDM). Receiver operating characteristics (ROC) analyses were used to assess the performance of standard descending aortic diameter and VDM derived growth rate for predicting TBAD. Optimal cut-points in continuous variables were determined using the method of Liu at al.^28^ We performed a sensitivity analysis restricted to the subgroup of patients with fibrillin-1 gene mutation or a history of lens dislocation/subluxation. All statistical analyses were performed using STATA (StataCorp. 2023. Stata Statistical Software: Release 18. College Station, TX: StataCorp LLC). A two-tailed p-value of ≤0.05 was considered statistically significant.

## RESULTS

### Study Population

From the initial cohort of 117 patients with MFS who met the inclusion criteria, 12 individuals were excluded—6 due to inadequate image quality and 6 due to VDM misregistration (**Figure 2**). **Supplementary Table 1** summarizes the clinical characteristics of included and excluded patients. Thus, 105 participants were included in our primary analysis. The study encompassed a clinical follow-up period of 8.6 years (range, 2.3-19.5), with the interval of imaging surveillance period of 5.3 years (range, 2.0-18.3). Among these, 74 (70.5%) individuals had a history of root/ascending repair (ARR), of which 17 (16.2%) were due to a history of repaired type A aortic dissection (DeBakey type II). The majority 67 (63.8%) of patients were male, with a mean age of 40.1 (range: 18.4-72.5). Additionally, 66 (62.9%) patients reported a family history of MFS, 38 (36.2%) with a history of ectopia lentis and 44 (93.6%) of the 47 patients with prior genetic testing had a pathogenic variant in the fibrillin-1 gene.

**Figure 2:**
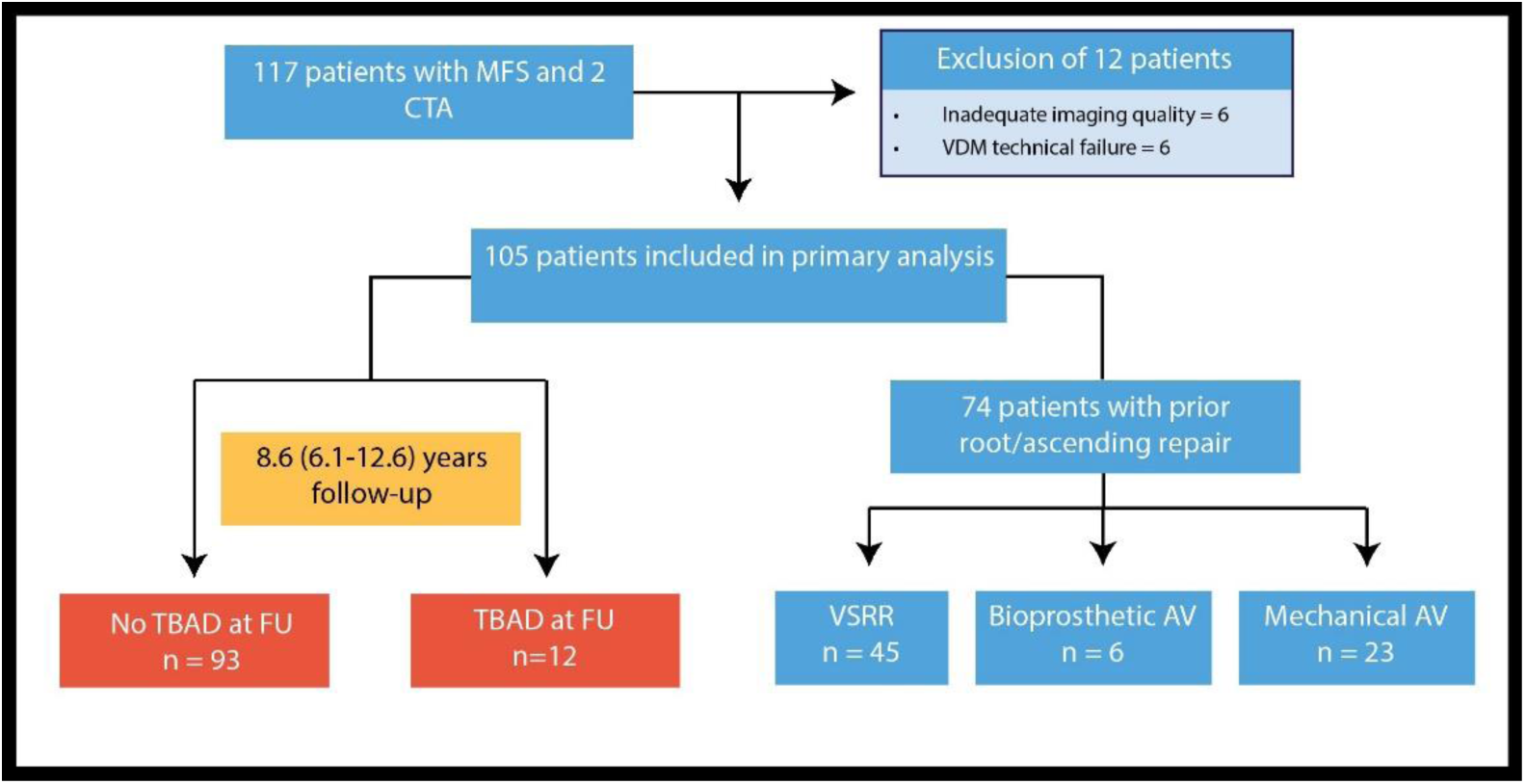
Study cohort overview. AVR = aortic valve. MFS = Marfan syndrome. VSRR = valve-sparing root replacement.

### Patient Characteristics and Aortic Metrics by Type B Aortic Dissection Status

#### Clinical characteristics

During the follow-up period, 12 out of 105 patients (11.4%) developed acute TBAD during imaging surveillance. Clinical features were largely similar between groups, except for a significantly higher proportion of females in the TBAD group compared to those without (TBAD: 66.7% vs no-TBAD 32.3%; *p* = 0.03), and a higher proportion of patients with aortic valve replacement compared to valve sparing repair (TBAD: 68.3% vs. no-TBAD: 27.3%; *p* = 0.02). Additionally, dural ectasia (TBAD: 91.7 vs no-TBAD: 43.5%; *p* = 0.002) and use of warfarin (TBAD: 58.3 vs no-TBAD: 24.7%; p = 0.04) was more frequent in patients with TBAD. The mean ages at first available CT [TBAD: 35.7 (IQR: 28.0-47.1) vs no-TBAD: 40.4 (IQR: 28.5-51.43) years, p=0.50], prior ascending repair (TBAD: 26.4 ± 3.3 vs no-TBAD: 31.5 ± 1.7 years, p=0.23) or onset of TBAD [TBAD: 41.4 (IQR: 34.9-53.5) vs no-TBAD: 49.4 (IQR: 37.9-62.2) years, p=0.18] did not significantly differ between groups. Clinical characteristics of patients by TBAD status are detailed in **Table 1**. When stratified by sex (**Supplementary Table 2)**, clinical characteristics were overall similar between female and male patients, except for a significantly higher proportion of dural ectasia in females (Females: 73.7 vs Males: 34.3 %; p < 0.001).

**Table 1:** Patient characteristics by TBAD status. Continuous variables values are mean ± SD or median (interquartile range). Binary variable values are n (%). ACE-I = angiotensin-converting enzyme inhibitors. ARB = angiotensin receptor blockers. COPD = chronic obstructive pulmonary disease. FU = follow-up. LDL = low-density lipoproteins. MFS = Marfan syndrome. TBAD = type B aortic dissection. VSRR = valve-sparing root replacement.

| Characteristics | No TBAD at FU<br>(n=93) | TBAD at FU<br>(n=12) | p |
| --- | --- | --- | --- |
| Age at first CT (years) | 40.4 (28.4-51.4) | 35.7 (28.0-47.1) | 0.50 |
| Sex (female) | 30 (32.3) | 8 (66.7) | **0.03** |
| Body mass index (kg/m<sup>2</sup>) | 24.8 (21.4-30.2) | 23.2 (20.7-27.5) | 0.37 |
| Diabetes | 8 (8.6) | 0 (0.0) | 0.59 |
| COPD | 6 (6.5) | 1 (8.3) | 0.58 |
| LDL at baseline (mg/dL) | 103.4 ± 29.0 | 70.0 ± 21.1 | **0.02** |
| LDL at second CT (mg/dL) | 96.1 ± 32.9 | 87.9 ± 9.0 | 0.50 |
| Coronary calcification | 35 (37.6) | 5 (41.7) | 0.76 |
| Aortic calcification | 53 (57.0) | 10 (83.3) | 0.12 |
| Chronic renal disease | 11 (11.8) | 0 (0) | 0.36 |
| Myocardial infarction | 4 (4.3) | 2 (16.7) | 0.14 |
| Heart failure | 15 (16.1) | 2 (16.7) | 1.00 |
| Hypertension | 36 (38.7) | 6 (50.0) | 0.54 |
| Systolic blood pressure (mmHg) | 120.8 ± 17.4 | 119.8 ± 16.9 | 0.85 |
| Diastolic blood pressure (mmHg) | 68.6 ± 10.7 | 65.7 ± 12.6 | 0.40 |
| Any anti-hypertensive | 76 (81.7) | 11 (91.7) | 0.69 |
| Beta-blockers | 71 (76.3) | 10 (83.3) | 0.73 |
| ARB | 30 (32.3) | 4 (33.3) | 1.00 |
| ACE-I | 6 (6.5) | 0 (0.0) | 1.00 |
| Calcium channel blockers | 7 (7.5) | 1 (8.3) | 1.00 |
| Diuretics | 14 (15.1) | 2 (16.7) | 1.00 |
| Statin use | 26 (28.0) | 6 (50.0) | 0.18 |
| Warfarin | 23 (24.7) | 7 (58.3) | **0.04** |
| Tobacco history | 27 (29.0) | 6 (50.0) | 0.19 |
| Cocaine use | 3 (3.2) | 0 (0.0) | 1.00 |
| Bicuspid aortic valve | 6 (6.5) | 1 (8.3) | 0.58 |
| Aortic stenosis | 1 (1.1) | 0 (0.0) | 1.00 |
| Aortic regurgitation | 4 (4.3) | 0 (0.0) | 1.00 |
| History of mitral valve prolapse | 47 (50.5) | 7 (58.3) | 0.76 |
| Mitral regurgitation | 9 (9.7) | 3 (25.0) | 0.14 |
| Positive family history of MFS | 59 (63.4) | 7 (58.3) | 0.76 |
| Fibrillin-1 pathogenic variant | 39 (92.9) | 5 (100) | 1.00 |
| Ectopia lentis | 31 (33.3) | 7 (58.3) | 0.11 |
| Dural ectasia | 40 (43.5) | 11 (91.7) | **0.002** |
| History of type A dissection | 13 (14.0) | 4 (33.3) | 0.10 |
| Prior ascending repair | 63 (67.7) | 11 (91.7) | 0.10 |
| - Age at repair | 31.5 ± 1.7 | 26.4 ± 3.3 | 0.23 |
| - VSRR | 42 (66.7) | 3 (27.3) | **0.02** |
| Cardiovascular death | 4 (4.3) | 4 (33.3) | **0.01** |
| All-cause death | 12 (12.9) | 5 (41.7) | **0.02** |
| Imaging surveillance interval | 5.2 (3.5-8) | 5.4 (4.1-7) | 0.94 |
| Clinical follow-up interval | 9.4 (6.3-12.8) | 6.9 (4.5-8.2) | **0.03** |

Of the 105 patients, 31 (30%) had a native ascending aorta, 45 (43%) had VSRR, 6 (6%) with bioprosthetic aortic valve replacement (AVR), 23 (22%) with mechanical AVR; clinical characteristics by valve type are described in **Supplementary Tables 3 and 4.** Patients with bioprosthetic AVR were significantly older than patients with prior VSRR (Bioprosthetic AVR: 58.2 vs VSRR: 38.0 years, *p =* 0.01) and native ascending aorta (Bioprosthetic AVR: 58.2 vs Native ascending aorta: 37.6 years, p = 0.01). Patients with mechanical AVR had higher frequency of aortic calcification, ectopia lentis, warfarin use and history of repaired DeBakey II type A dissection. On univariate analysis, TBAD frequency was higher in mechanical AVR patients compared to VSRR (Mechanical AVR: 34.8 vs VSRR: 3.7 %; p = 0.01) and native roots (Mechanical AVR: 34.8 vs VSRR: 3.2 %; *p* = 0.003).

In 8 out of 12 cases, TBAD marked the patient’s first aortic dissection, whereas 4 had a prior history of repaired Debakey type II dissection. Cardiovascular mortality at follow-up was higher in patients with TBAD compared to those without (TBAD: 33.3 vs no-TBAD: 4.3%; *p* = 0.01). A total of 17 patients died during the follow-up period, with 3 attributed to aortic causes, 5 due to non-aortic cardiovascular disease, 2 due to non-cardiovascular causes (malignancy) and 7 due to unknown causes.

#### Aortic Diameters at Baseline and Follow-up Imaging

**Table 2** summarizes the anatomic imaging biomarkers of patients with and without type B aortic dissection. In the pre-dissection imaging of the 12 patients who developed type B dissection, the average diameter of the corresponding segment where the entry tear developed was 32.8 ± 5.9 mm. Entry tears were located in the proximal segment of the descending thoracic aorta in all 12 TBAD cases, with the maximum diameter of the descending aorta located in a different segment in 7 cases (58%). Among TBAD cases, the descending corresponding segment was normal (<3.0 cm) in 3 (25%), mildly dilated (range: 3.0-3.9 cm) in 7 (58%), and significantly enlarged (>= 4.0 cm) in 2 (17%). Baseline aortic diameters at the distal descending (TBAD: 26.1 vs no-TBAD: 22.0 mm; *p* = 0.01), celiac artery (TBAD: 27.5 vs no-TBAD: 22.7 mm; *p* = 0.01), and superior renal artery (TBAD: 23 vs no-TBAD: 21.8 mm; *p* = 0.02) were larger patients with TBAD. At follow-up, all segments of the thoracoabdominal aorta were significantly larger in those that developed TBAD, except for at the mid descending level (TBAD: 27.2 vs no-TBAD: 24 mm; *p* = 0.07). Change in maximal diameter of the descending aorta by standard diameter measurement technique was also significantly higher in patients with TBAD (0.88 vs 0.36 mm/y; *p* = 0.01). The optimal cut-point between TBAD and non-TBAD groups based on maximal descending diameter was 31.1 mm, which yielded a sensitivity of 75.0 % (95% CI 42.8-94.5%), specificity of 78.5 % (95% CI 68.8-86.3%), and area under the curve (AUC) of 0.77 (95% CI 0.63-0.90). When using change of maximal descending diameter, the sensitivity was of 83.3 % (95% CI 51.6-97.9%), specificity of 68.8 % (95% CI 58.4-78.0%), and area under the curve (AUC) of 0.77 (95% CI 0.63-0.90). No differences in aortic diameters were seen when stratifying by sex (**Supplementary Table 5).**

**Table 2:** Baseline and follow-up diameters of patients without and with TBAD. Values are mean ± SD or median (quartile 1-quartile 3). TBAD = type B aortic dissection. FU = follow-up.

| Aortic diameters (mm) | Baseline |  |  | Follow-up |  |  |
| --- | --- | --- | --- | --- | --- | --- |
|  | No TBAD at FU<br>(n=93) | TBAD at FU<br>(n=12) | p | No TBAD at<br>FU (n=93) | TBAD at FU<br>(n=12) | p |
| Proximal descending | 26.4 ± 3.6 | 27.7 ± 15.2 | 0.25 | 28.0 (25.8-30.5) | 29.8 (27.3-32.8) | 0.047 |
| Mid descending | 23.4 (21.0-26.0) | 26 (22.4-29.5) | 0.14 | 24.0 (22.0-27.0) | 27.2 (23.5-33.0) | 0.07 |
| Distal descending | 22.0 (20.0-25.0) | 26.1 (22.5-28.1) | 0.01 | 24.0 (21.7-26.6) | 32.8 (23.6-38.3) | 0.002 |
| Proximal to the celiac trunk | 22.7 (21.0-26.0) | 27.5 (23.8-32.0) | 0.01 | 24.6 (22.0-27.2) | 34.8 (25.1-41.0) | 0.001 |
| Proximal to the mesenteric artery | 22.0 (20.1-24.5) | 24.0 (22.0-26.9) | 0.06 | 23.3 (21.4-25.8) | 28.8 (25.0-36.1) | <0.001 |
| Proximal to the superior renal artery | 21.8 (19.0-24.0) | 23.0 (22.3-25.0) | 0.02 | 22.8 (20.0-24.2) | 28.0 (24.0-35.0) | <0.001 |
| Distal to the inferior renal artery | 18.5 (16.6-21.0) | 21.0 (20.0-21.0) | 0.09 | 20.0 (18.0-22.0) | 24.0 (20.5-25.0) | 0.003 |
| 15 mm distal to the inferior renal artery | 18.0 (16.4-19.9) | 21.0 (16.0-22.0) | 0.39 | 19.6 (17.5-21.4) | 22.2 (21.7-24.6) | 0.01 |
| Maximal thoracic descending | 27.0 ± 4.0 | 29.4 ± 1.31 | 0.06 | 28.5 (26.0-30.9) | 33.2 (30.1-38.8) | 0.001 |

#### Elongation, Arch Angle and Tortuosity Index

Baseline values of arch angle, descending length and tortuosity index and their change over follow-up are detailed in **Supplementary Table 6 and Table 3**. Of note, 11 patients were excluded from aortic length analyses due to the lack of imaging coverage to the level of the celiac artery. Patients with type B dissection had significantly greater descending length corrected for height at follow-up (TBAD: 138.6 vs no-TBAD: 128.5 mm/m, *p* = 0.02). Elongation rate in descending thoracic aorta was also significantly greater in those with vs without type B dissection (TBAD: 2.4 vs no-TBAD: 0.5 mm/y; *p* < 0.001). **Supplementary Figure 4** depicts a patient with significant elongation over time (18.8 mm over 5 years). Elongation rate was weakly correlated with rate of growth in the radial direction by VDM in the overall cohort (r=0.31, p=0.002).

Baseline aortic arch angle was significantly less acute in TBAD group (TBAD: 90° vs. no-TBAD: 82°, *p*=0.049), however, TBAD patients demonstrated increasing acuity of arch angle over follow-up (TBAD: -5.2° vs. no-TBAD: +2.1°, *p* = 0.03). Tortuosity index of the descending thoracic aorta was not significantly different at baseline or follow-up CTAs, however, there was a trend towards increased tortuosity in the TBAD group over follow-up (TBAD: +0.04 vs. no-TBAD: +0.02, p=0.08). Sex-differences in elongation, arch angle, and tortuosity index were not observed, and no differences in descending aortic lengths were seen when corrected for height (**Supplementary Table 7 and 8).**

#### Radial growth (VDM)

**Table 3** and **Figure 3** summarizes the radial growth rate assessed by VDM in different segments of patients with and without type B aortic dissection. When assessed by VDM, radial growth across descending aortic segments was significantly greater in patients with TBAD both in the overall descending thoracic aorta (TBAD: 0.63 vs no-TBAD: 0.23 mm/y, *p* < 0.001) and when examined by proximal and distal descending segments separately. Representative VDM assessments of 3 TBAD patients with native root (**Figure 4**), repaired root and diffuse native aortic growth (**Figure 5**), and repaired root with focal distal arch growth (**Figure 6**) are shown. Among the 35 patients in the upper tertile of radial growth rate demonstrated a growth rate (>0.35 mm/y) the vast majority (29/35, 82.9%) exhibited diffuse growth, while 6 (17.1%) showed growth localized to the proximal descending aorta. The majority (10/12, 83%) of patients with TBAD demonstrated diffuse descending aortic growth. The optimal cut-point in radial growth rate for the outcome of TBAD was 0.4 mm/y, with 11 out of 12 (91.7%) having a VDM derived growth above this threshold and 19 out of 93 (20.4%) in the no-TBAD below this threshold, yielding a sensitivity of 91.7% (95% CI 61.5-99.8%), specificity of 79.6% and AUC of 0.86 (95% CI 0.77-0.95). Sensitivity analysis restricted to patients with pathogenic/likely pathogenic variant in fibrillin-1 gene or a history of ectopia lentis yielded similar results (**Supplementary Table 9**). When stratified by sex, no significant differences were seen in growth rate by VDM (**Supplementary Table 10**).

**Table 3:** Change in centerline-derived imaging biomarkers of patients without and with TBAD. Values are mean ± SD or median (quartile 1-quartile 3). TBAD = type B aortic dissection. FU = follow-up. *82 for length analyses.

| Change in centerline-derived imaging biomarkers | No TBAD at FU (n=93)* | TBAD at FU (n=12) |  |
| --- | --- | --- | --- |
| Elongation (mm) | 2.0 (-0.9, 8.0) | 11.3 (5.3-19.3) | 0.002 |
| Elongation rate (mm/y) | 0.5 (-0.2, 1.4) | 2.4 (1.3-4.5) | <0.001 |
| Change in descending tortuosity index | +0.02 (0-0.04) | +0.04 (0.02-0.07) | 0.08 |
| Aortic arch angle change (°) | 5.2 ± 10.3 | -2.1 ± 14.7 | 0.03 |

**Figure 3:**
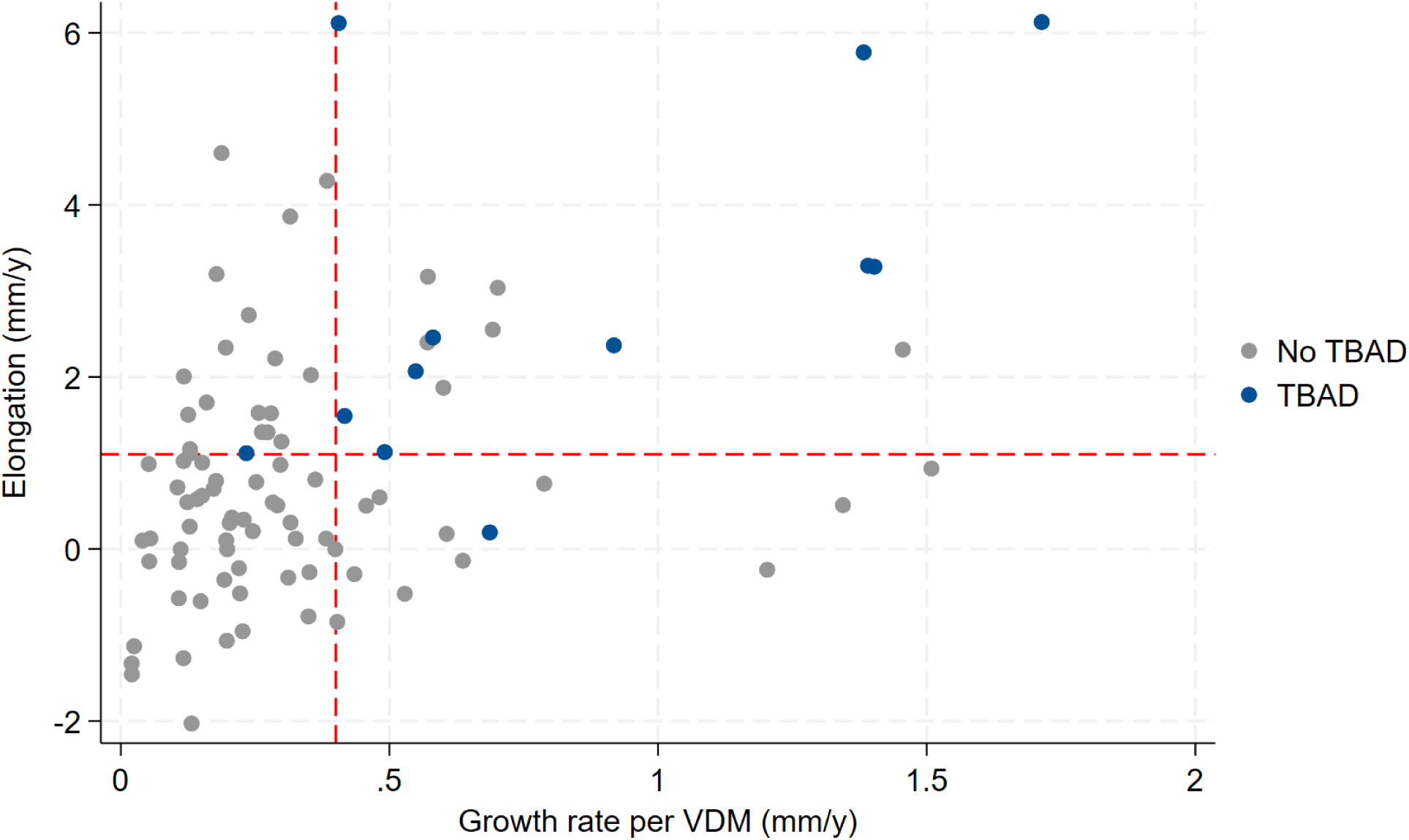
Scatter plot of elongation vs. growth rate per VDM for patients with and without TBAD. Scatter plot illustrating the relationship between elongation (mm/year) on the y-axis and growth rate per VDM (mm/year) on the x-axis for patients with Marfan syndrome, comparing those without TBAD (gray dots) against those with TBAD (blue dots). The horizontal red dashed line represents the optimal cut point for elongation (1.1 mm/y), while the red vertical dashed line signifies the optimal cut point for growth rate per VDM (0.4 mm/y). Note: Two outliers with high negative elongation due to spine disease were excluded from visualization.

**Figure 4:**
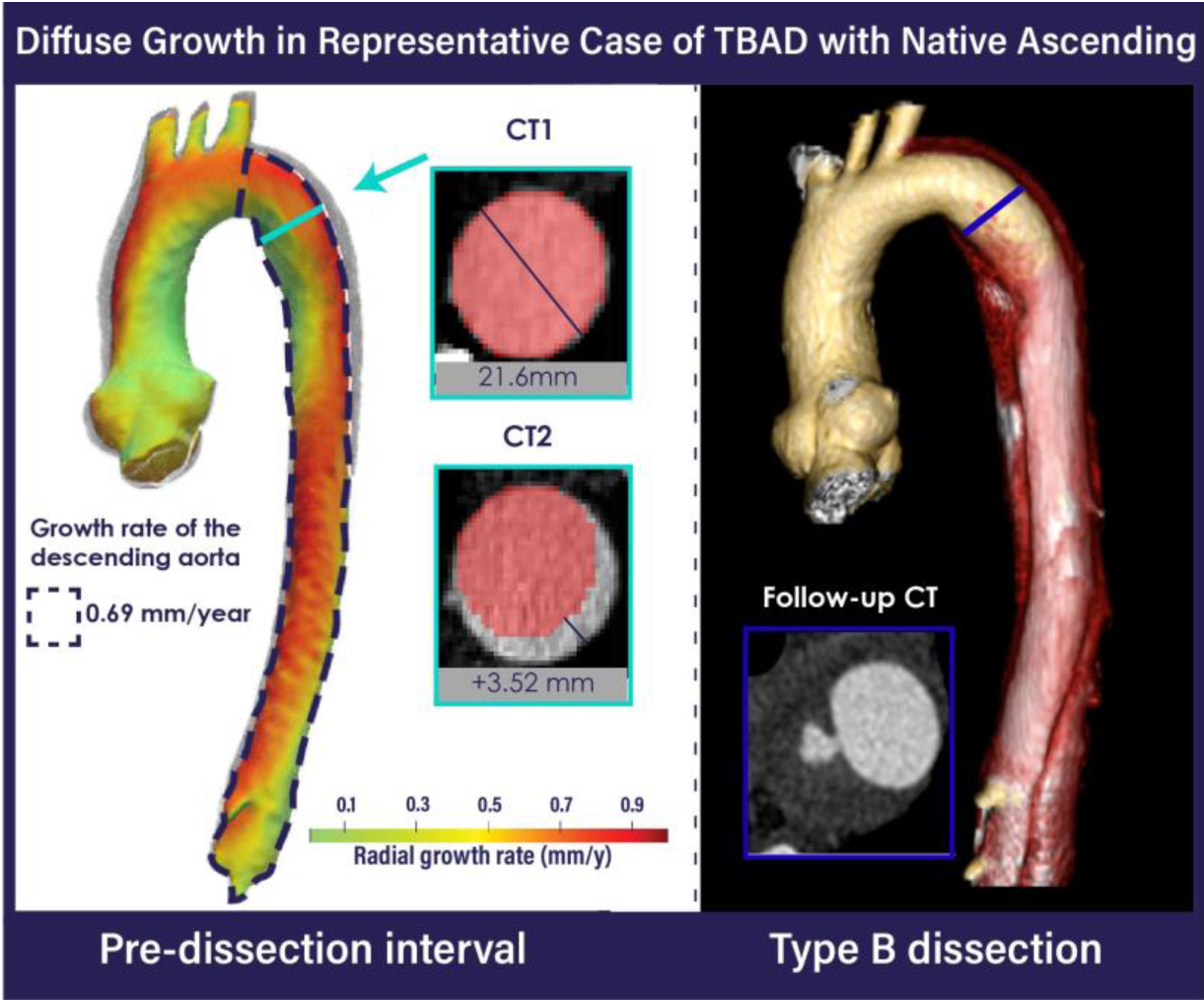
Representative VDM analysis of a patient with MFS and native ascending thoracic aorta, demonstrating diffuse growth. Red masks depicting the baseline anatomy of the proximal descending are overlaid on both CT scans used for VDM analysis. Dashed lines indicate the area of the descending thoracic aorta used for growth extraction from VDM. The 3D model and axial plane view on the right demonstrate type B dissection at follow-up.

**Figure 5:**
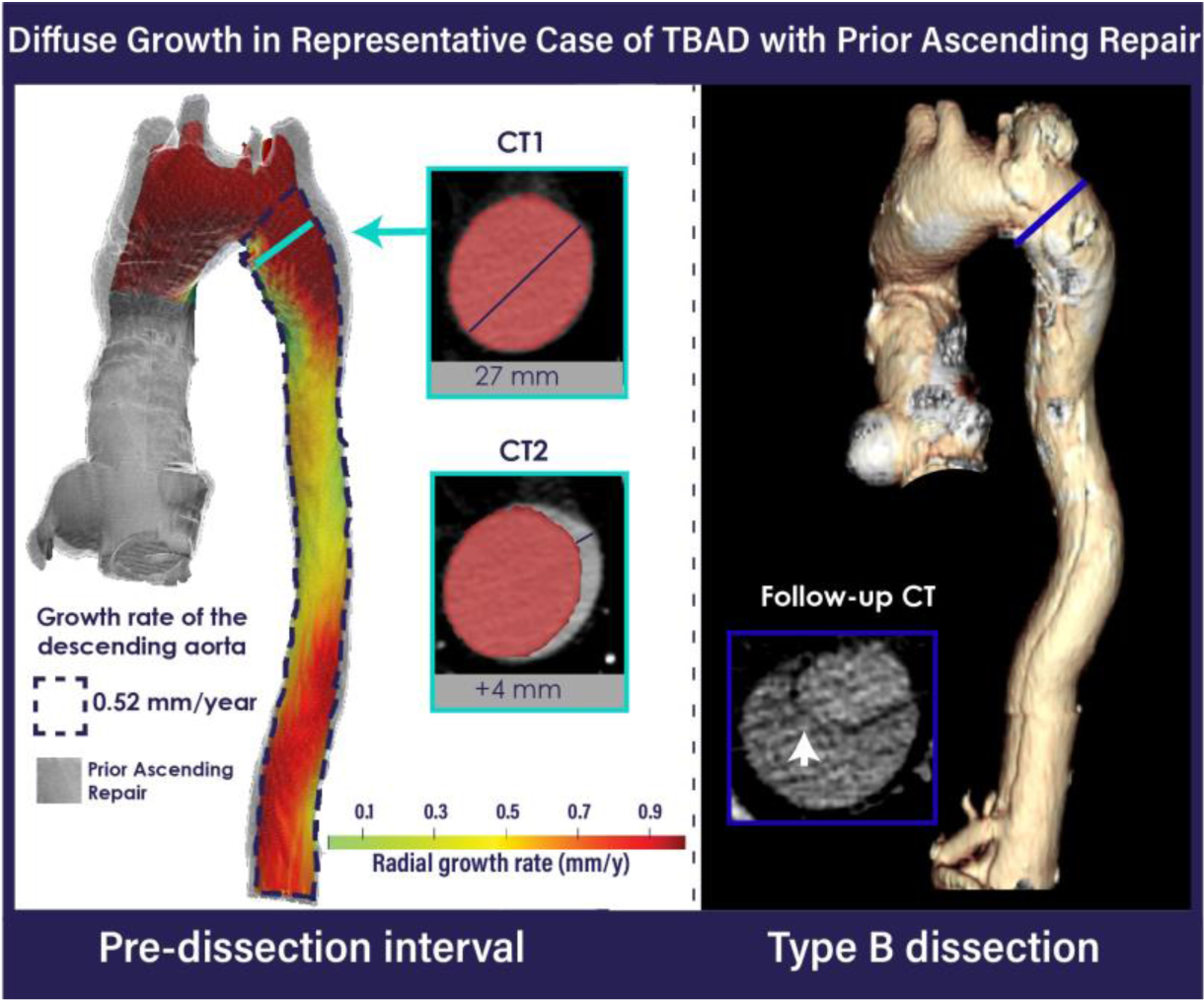
Representative VDM analysis of a patient with Marfan syndrome with prior root and ascending repair (gray region), demonstrating diffuse growth of the aortic arch and descending. Red masks depicting the baseline anatomy of the proximal descending are overlaid on CT scans used for VDM analysis. Dashed lines indicate the area of the descending thoracic aorta used for growth extraction from VDM. The 3D model and intimal tear (white arrow) on axial view on the right demonstrate type B dissection at follow-up.

**Figure 6:**
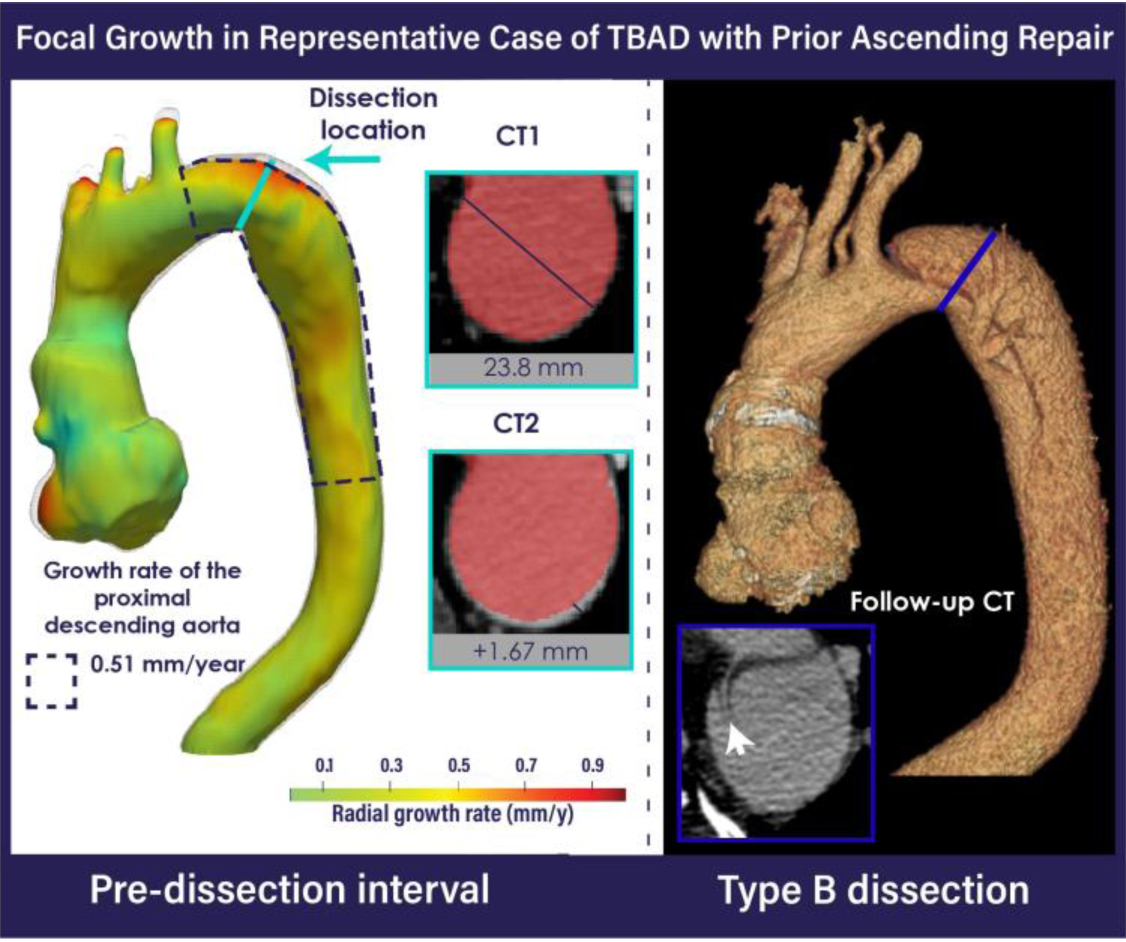
Representative VDM analysis of a patient with MFS and prior root and ascending repair (gray zone), demonstrating focal growth on the proximal descending. Red masks depicting the baseline anatomy of the proximal descending are overlaid on both CT scans used for VDM analysis. Dashed lines indicate the area of the descending thoracic aorta used for growth extraction from VDM. The 3D model and tear (white arrow) on axial view on the right demonstrate type B dissection at follow-up.

#### Sex differences

Baseline clinical characteristics and aortic imaging biomarkers by sex are detailed in **Supplementary Tables 5-8.** Clinical characteristics were overall similar between female and male patients, except for a significantly higher proportion of dural ectasia in females (Females: 73.7 vs Males: 34.3 %; *p* < 0.001). Sex-differences in descending aortic lengths were not seen when corrected for height. No significant differences were seen in growth rate by VDM or other imaging biomarkers.

#### Thoracoabdominal calcification

Baseline clinical characteristics and aortic imaging biomarkers by presence or absence of thoracoabdominal calcification (TAC) are detailed in **Supplementary Tables 11, 12, 13, 14 and 15.** Patients with TAC were significantly older (TAC: 46.8 vs no-TAC: 29.5 years; p < 0.001) and were more likely to be using diuretics (TAC: 23.8 vs no-TAC: 2.4 %; *p* = 0.002), statin (TAC: 41.3 vs no-TAC: 14.3 %; *p* = 0.01), and warfarin (TAC: 44.4 vs no-TAC: 4.8 %; *p* < 0.001). No difference was seen in LDL levels in closest to CT1 or CT2 (TAC: 100.9 vs no-TAC: 101.0 mg/dL; *p* = 0.98 and TAC: 90.4 vs no-TAC: 105.8 mg/dL; *p* = 0.09, respectively). Patients with TAC had higher cardiovascular death and overall mortality events on univariate analysis (TAC: 12.7 vs no-TAC: 0.0 %; *p* = 0.02 and TAC: 23.8 vs no-TAC: 4.8 %; *p* = 0.01, respectively). After adjustment for age, the presence of TAC conferred high risk of TBAD (OR = 7.8, 1.3-47.7, 95% CI). Further, patients with TAC exhibited higher elongation rate (TAC: 1.02 vs no-TAC: 0.26 mm/y; *p* = 0.003), but there was no difference in VDM derived radial growth rate (TAC: 0.25 vs no-TAC: 0.27 mm/y; *p* = 0.98).

#### Aortic Metrics by Ascending Repair Status

Baseline clinical characteristics and aortic imaging biomarkers between patients with previous ARR and those with a native ascending aorta are detailed in **Supplementary Tables 16, 17, 18, 19 and 20**. Patients with previous ARR exhibited longer length of the descending aorta at baseline and follow-up and larger thoracoabdominal aortic diameters. No significant differences were seen in absolute and yearly changes in length, arch angle or tortuosity metrics by ARR status. VDM derived growth rate of the descending thoracic aorta was significantly higher in patients with previous ARR compared to those with native ascending aorta (ARR: 0.29 vs Native ascending aorta: 0.18 mm/y, *p* = 0.001).

### Multivariable Predictors of Type B Aortic Dissection and Growth Rate

Multivariable logistic regression analysis revealed an increased risk of TBAD with increasing radial growth rate (OR 9.78, 95% CI 1.11-86.08; *p* = 0.04) and elongation rate (OR 2.16, 95% CI 1.28-3.65; *p* = 0.004), but not with age at ARR, female sex, and pre-dissection proximal descending aortic diameter (**Table 4)**. When examining predictors of VDM-derived radial growth rate (log_10_), multivariable linear regression identified positive associations with maximum diameter of the descending aorta at baseline (β = 0.099, 95% CI 0.054-0.144; *p* < 0.001), history of tobacco (β = 0.326, 95% CI 0.001-0.652; *p* = 0.049) and use of warfarin (β = 0.492, 95% CI 0.117-0.867; *p* = 0.01), as well as a negative association with age (β = -0.034, 95% CI -0.047, -0.021; *p* < 0.001). No independent associations of radial growth rate were seen with sex, history of ARR, dural ectasia, or TAC (**Table 5)**.

**Table 4:**
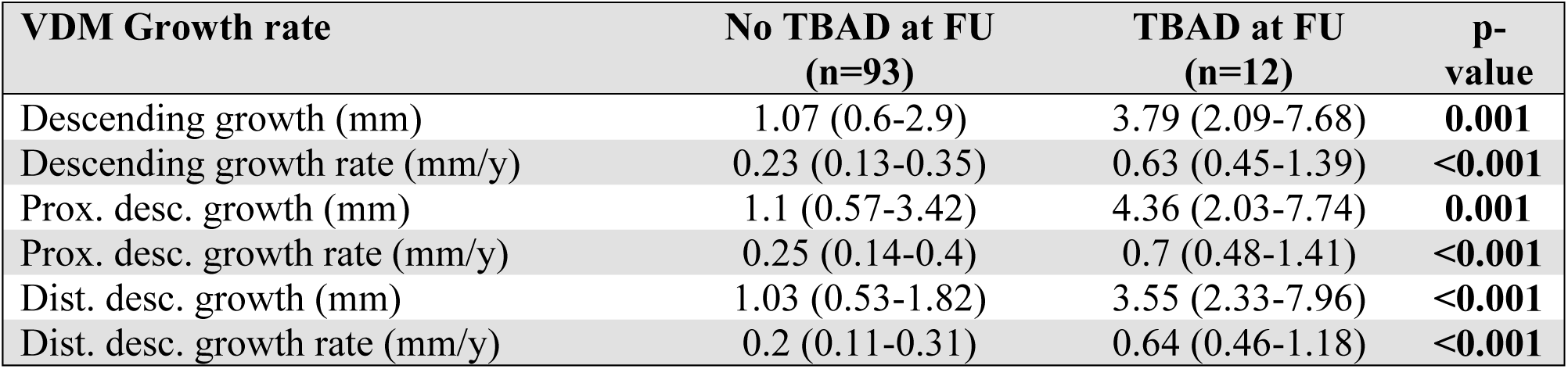
Radial growth assessed by VDM of patients without and with TBAD. Values are mean ± SD or median (quartile 1-quartile 3). Bold p values are statistically significant. TBAD = type B aortic dissection. FU = follow-up. VDM = Vascular Deformation Mapping.

| VDM Growth rate | No TBAD at FU<br>(n=93) | TBAD at FU<br>(n=12) | p-value |
| --- | --- | --- | --- |
| Descending growth (mm) | 1.07 (0.6-2.9) | 3.79 (2.09-7.68) | **0.001** |
| Descending growth rate (mm/y) | 0.23 (0.13-0.35) | 0.63 (0.45-1.39) | **<0.001** |
| Prox. desc. growth (mm) | 1.1 (0.57-3.42) | 4.36 (2.03-7.74) | **0.001** |
| Prox. desc. growth rate (mm/y) | 0.25 (0.14-0.4) | 0.7 (0.48-1.41) | **<0.001** |
| Dist. desc. growth (mm) | 1.03 (0.53-1.82) | 3.55 (2.33-7.96) | **<0.001** |
| Dist. desc. growth rate (mm/y) | 0.2 (0.11-0.31) | 0.64 (0.46-1.18) | **<0.001** |

**Table 5:** Logistic regression for TBAD. Bold p values are statistically significant. FU = follow-up. TBAD = type B aortic dissection. VDM = Vascular Deformation Mapping.

| Variables | OR | 95% CI | p-value |
| --- | --- | --- | --- |
| Age at repair (per year) | 1.02 | 0.94-1.12 | 0.58 |
| Female sex | 2.68 | 0.41-17.68 | 0.31 |
| Pre-dissection proximal descending aortic diameter (per mm) | 0.99 | 0.82-1.21 | 0.94 |
| VDM radial growth rate (per mm/y) | 9.78 | 1.11-86.08 | **0.04** |
| Elongation rate (per mm/y) | 2.16 | 1.28-3.65 | **0.004** |

**Table 6:** Multivariate linear regression for VDM derived growth rate. Bold p values are statistically significant. Interpretation for beta coefficients: per one-unit increase of the **dependable** variable, growth rate will be multiplied by 10^beta^; e.g.: per 1 mm (one-unit) increase in proximal diameter, the expected increase in growth rate is 10^0.099^ ∴ 1.258, 25.8% increase. FU = follow-up. TBAD = type B aortic dissection. VDM = Vascular Deformation Mapping.

| Variables | Growth rate of descending thoracic aorta<br>(Log<sub>10</sub>) |  |  |
| --- | --- | --- | --- |
| | $\beta$ | 95% CI | p-value |
| Age | -0.034 | -0.047, -0.021 | **<0.001** |
| Female sex | -0.060 | -0.398, 0.278 | 0.72 |
| History of aortic root repair | 0.356 | -0.003, 0.714 | 0.052 |
| Maximum diameter of the descending aorta at baseline | 0.099 | 0.054-0.144 | **<0.001** |
| Tobacco | 0.326 | 0.001-0.652 | **.049** |
| Dural ectasia | 0.179 | -0.150, 0.508 | 0.28 |
| Thoracoabdominal aortic calcification | -0.161 | -0.546, 0.223 | 0.41 |
| Warfarin | 0.492 | 0.117-0.867 | **0.01** |
| Beta-blockers | -0.157 | -0.598, 0.217 | 0.41 |
| Angiotensin-converting enzyme inhibitors | -0.268 | -0.598, 0.063 | 0.11 |

## DISCUSSION

The results of this study revealed several unique and clinically relevant insights regarding imaging biomarkers that may confer risk of TBAD among patients with Marfan syndrome. First, our findings confirm prior reports showing the TBAD in MFS tends to occur in non-dilated or minimally dilated segments, at sizes far below typical surgical repair thresholds. Second, we demonstrate that both radial growth rate, derived from VDM, and elongation rate of the descending thoracic aorta, derived from centerline measurement, are independent predictors of TBAD development. Third, TBAD patients demonstrated higher pre-dissection diameters in their distal thoracic and abdominal aortic segments suggesting a more diffuse and severe aortic disease phenotype. Although underpowered for multivariable analysis, we identified high rates of dural ectasia, warfarin use, and mechanical aortic valve replacement in our TBAD group, suggesting that these factors may be associated with risk of dissection, either directly or via associations with a more severe disease phenotype. Fourth, we identified younger age, baseline maximal diameter of the descending thoracoabdominal aorta, smoking history, and use of warfarin to be independent predictors of aortic growth rate in MFS. Lastly, patients with atherosclerotic calcification of the thoracoabdominal aorta displayed a higher risk for TBAD after age adjustment, as well as a higher rate of descending aortic elongation, although not radial growth rate.

### Diameter and other anatomic parameters

Patients in our cohort with TBAD exhibited greater degrees of thoracoabdominal aortic dilation on pre-dissection imaging, specifically with larger diameters in both supra- and infra-renal aortic segments, suggesting a more diffuse aortic involvement. This finding aligns with previous reports demonstrating that MFS patients with TBAD at follow-up exhibit increased abdominal aortic diameters before the event.^6^ A diffuse disease phenotype has also been described beyond the aorta, with aortic branch aneurysms being present in one-quarter of patients with MFS, associated with older age and greater degrees of root dilation, and independently predicted need for aortic surgery.^29^

Increased aortic length and tortuosity have been reported in patients with TAAD and TBAD when compared to aneurysmal or control populations; however, our study is the first to show that elongation rate of the descending thoracic aorta is an independent risk factor for TBAD in MFS.^30–33^ Ascending aortic elongation rate has been posited as an underappreciated metric of disease progression, although prior to this study there has been a lack of data linking elongation rate to development of dissection.^34^ Our findings are in agreement with prior studies that higher degrees of tortuosity – a result of elongation – increases risk of adverse cardiovascular events in patients with genetic aortopathy.^35^

### Growth rate assessed by VDM

A key finding of our study is that radial growth rate of the descending thoracic aorta by VDM was an independent predictor of TBAD dissection during follow-up. Specifically, we identified that a growth rate threshold of ≥0.4 mm/year conferred a high sensitivity (92%) and specificity (80%) for discriminating TBAD patients, a subgroup in which medical therapy could be pursued most aggressively. Comparing the performance of VDM growth (≥0.4 mm/year) and change in maximal diameter of the descending aorta by the standard diameter measurement technique (>31 mm), there was a moderate improvement in both sensitivity (92% vs. 83%, respectively) and specificity (80% vs. 69%, respectively). Additionally, VDM analysis offers the advantage of being 3D, thus not dependent on a single measurement plane/location. The potential negative impact of variability in diameter measurement location was evident in our data, with sensitivity dropping to 58% when the diameter of the proximal descending (i.e., the location of entry tear formation) was used in place of the maximal diameter to calculate growth by clinical CT.

The link between fast growth and risk of dissection in MFS has been previously reported by den Hertog et al., who reported a similar growth rate cut point (i.e., ≥0.5 mm/year) to our growth rate per clinical CT and per VDM (0.56 and 0.4 mm/year, respectively).^6^ However, den Hertog et al showed greater overlap in aortic growth rates between groups with a lower AUC for accelerated growth compared to our study (e.g., 0.69 vs. 0.86). Improved growth rate prediction of TBAD in our study may be in part due to the improved accuracy and comprehensiveness of growth measurement by the VDM technique compared standard diameter-based measurement techniques;^23^ an advantage that may be critical for identifying fast growth over shorter intervals (e.g., a growth rate of 0.5 mm/year would result in only 1.5 mm of growth over 3 years). Additionally, we observed both localized and diffuse growth patterns in MFS patients, further emphasizing the benefit of a three-dimensional growth assessment to more clearly depict the extent of growth, especially growth occurring outside of standard measurement locations or regions of dilation. While not the focus of this study, assessment of unique growth patterns by VDM in combination with other genetic and serologic markers may greatly advance personalized disease phenotyping in MFS.

### Age and Sex differences

We did not find an association between age (at ARR or onset of TBAD) and the occurrence of TBAD on univariate or multivariable analyses. However, we did observe an association between younger age and faster descending aortic growth on multivariable analysis. Our findings agree with den Hartog et al. who also found no association between age and TBAD occurrence, however, a recent report from the Cornell Aortic Aneurysm Registry suggested that older age was associated with risk of TBAD on multivariable analysis.^6, 36^

While we did observe a higher proportion of females with TBAD in our study, although after correcting aortic dimensions for body size and adjusting for co-variates, we did not identify any sex-specific associations with aortic growth or TBAD occurrence. The GenTAC registry reported a higher rate of aortic interventions in males with MFS than females, but these differences were similarly not seen after correcting for body size.^37^ Of note, we observed a higher incidence of dural ectasia in women (74% in women vs. 34% in men), which may explain the overrepresentation of women with TBAD given that dural ectasia was associated with TBAD in our univariate – although not multivariate – analysis and in a recent report from the Cornell Aortic Aneurysm Registry.^36^ While the exact mechanism remains unclear, the concept that dural ectasia may mark a more severe phenotype is supported by mouse studies of MFS showing severe abnormalities in dural structure and elastic fiber composition.^38^

### Repaired vs Native Ascending

Despite 11 out of 12 patients in the TBAD group having undergone prior ascending repair, we observed no statically significant difference in the incidence of TBAD with prior ascending aortic repair, unlike prior reports.^6, 11, 39^ This difference is likely explained by the low frequency of patients with native aortic roots (29%) in our CT-based study cohort, due to a preference for MRI and echocardiography surveillance in this group.^6, 11, 39^ Previous research has highlighted post-operative abnormalities in aortic arch angulation, blood flow dynamics (i.e., wall shear stress) and pulsatile loading of the distal native aorta following ascending repair that may predispose to TBAD.^40–42^ While further research examining the relationships between anatomic and biomechanical factors is needed to more clearly understand the pathogenesis of TBAD in MFS, our study strongly suggests that aortic growth rate is an important factor to incorporate in such studies.

### Warfarin Use and Thoracoabdominal Calcification

Warfarin use predicted descending aortic growth rate in our study despite adjustment for patient factors such as age, aortic diameter, and sex. This observation may in part reflect a more severe disease phenotype via its association with mechanical aortic valve replacement. However, warfarin itself has a variety of vitamin-K related effects on aortic wall biology including effects on smooth muscle cell phenotype and pro-inflammatory effects which could promote aortic growth and aneurysm formation independent of valve replacement.^43^

The role of warfarin in promoting vascular calcification has been well-described^44^ and these effects may partially explain the observation that 60% of our MFS cohort demonstrated aortic calcification. However, only 44% of MFS patients with aortic calcification had a history of warfarin use, and the rates of aortic calcification in our relatively young MFS cohort (mean age of 40 years) were comparable to large population studies with average age of approximately 60 years.^45^ Studies in both surgical aortic tissue and murine aortic smooth muscle cells have also shown that elastin fragmentation – a hallmark of MFS – may itself promote aortic micro-calcification by destabilizing extracellular matrix and promoting osteogenic smooth muscle phenotypes.^46^ The presence of aortic calcification in our study was associated with higher rates of elongation and TBAD after adjusting for age, findings which support the idea that aortic calcification (micro- and macro-) may be relevant in predicting aortic disease progression in adult MFS patients. As MFS patients live longer due to advances in medical and surgical therapy, new challenges may arise requiring further research related to the effects of traditional vascular risk factors such as obesity and atherosclerosis on this population’s intrinsically fragile aortic substrate.^47^

### Limitations

Despite the advantages of a retrospective study spanning 20 years of imaging and clinical follow-up to capture rare events such as TBAD, there are relevant variables which are difficult to accurately capture retrospectively (e.g., blood pressure control, detailed family history). Additionally, given the use of VDM analysis, our study was restricted to patients surveilled by CT, likely biasing our study cohort to older patients with high rates of ARR. Furthermore, given the challenge in disentangling highly correlated clinical characteristics (e.g., mechanical valve replacement, warfarin use and aortic calcification), the independent effects of such factors on outcomes remains unclear and should be examined in prospective studies. Finally, our analysis focused on a linearized growth rate over the longest available imaging internal to maximized growth detection, and we did not examine potential changes in growth rate over time and its associated risks; future efforts will examine imaging sub-intervals to better understand aortic growth trajectories and temporal relationships with TBAD.

## CONCLUSION

In this cohort of patients with MFS, radial growth and elongation rates of the descending thoracic aorta were independent predictors of TBAD occurrence during imaging surveillance. TBAD often occurred in non-dilated or slightly dilated descending thoracic aorta and was not predicted by pre-dissection aortic diameter or age at ARR. These findings provide important insights and mark an important step forward in predicting and managing aortic complications in MFS.

## Data Availability

The data that support the findings of this study are available from the corresponding author upon reasonable request.

## CLINICAL PERSPECTIVE

### What is new?

- Descending aortic radial growth by vascular deformation mapping (VDM) and elongation rates were independent predictors of TBAD occurrence during imaging surveillance.
- Among Marfan syndrome (MFS) patients that developed type B aortic dissection (TBAD), higher rates of dural ectasia, warfarin use, and mechanical aortic valve replacement were observed.
- TBAD commonly occurred in non-dilated or slightly dilated aortic segments and was not predicted by pre-dissection diameter or age at root replacement in multivariate analysis.

### What are the Clinical Implications?

- Additional features and biomarkers beyond aortic diameter can be used to assess aortic disease severity in patients with MFS, as aortic growth and elongation.
- VDM image analysis technique provides important insights regarding aortic growth diffuseness and severity in patients with MFS.
- Additional studies are needed to identify factors that contribute to the observed associations.

## Abbreviations and acronyms

3D: three-dimensional
AV: aortic valve
ARR: aortic root replacement
CTA: computed tomography angiography
ECG: electrocardiography
MFS: Marfan syndrome
TAC: thoracoabdominal aortic calcification
TAAD: type A aortic dissection
TBAD: type B aortic dissection
VDM: Vascular Deformation Mapping
VSRR: valve-sparing root replacement

## ACKNOWLEDGMENTS

We would like to acknowledge the Cardiovascular Health Improvement Project (CHIP) and MI-AORTA programs at the Frankel Cardiovascular Center for their support of this project through programmatic and database resources. We would like to acknowledge the Ahluwalia Singh Family Fund.

## SUPPLEMENTAL MATERIAL

Tables S1–S20

Figure S1-S4

## REFERENCES

1. Harris SL, Lindsay ME. Role of clinical genetic testing in the management of aortopathies. Current Cardiology Reports. 2021;23

2. Kwartler CS, Gong L, Chen J, Wang S, Kulmacz R, Duan XY, et al. Variants of unknown significance in genes associated with heritable thoracic aortic disease can be low penetrant “risk variants”. Am J Hum Genet. 2018;103:138–143

3. Silverman DI, Burton KJ, Gray J, Bosner MS, Kouchoukos NT, Roman MJ, et al. Life expectancy in the marfan syndrome. The American Journal of Cardiology. 1995;75:157–160

4. Finkbohner R, Johnston D, Crawford ES, Coselli J, Milewicz DM. Marfan syndrome. Long-term survival and complications after aortic aneurysm repair. Circulation. 1995;91:728–733

5. Gott VL, Greene PS, Alejo DE, Cameron DE, Naftel DC, Miller DC, et al. Replacement of the aortic root in patients with marfan’s syndrome. N Engl J Med. 1999;340:1307–1313

6. den Hartog AW, Franken R, Zwinderman AH, Timmermans J, Scholte AJ, van den Berg MP, et al. The risk for type b aortic dissection in marfan syndrome. J Am Coll Cardiol. 2015;65:246–254

7. Lenz A, Warncke M, Wright F, Weinrich JM, Schoennagel BP, Henes FO, et al. Longitudinal follow-up by mr angiography reveals progressive dilatation of the distal aorta after aortic root replacement in marfan syndrome. Eur Radiol. 2023

8. de Beaufort HWL, Trimarchi S, Korach A, Di Eusanio M, Gilon D, Montgomery DG, et al. Aortic dissection in patients with marfan syndrome based on the irad data. Ann Cardiothorac Surg. 2017;6:633–641

9. Groth KA, Stochholm K, Hove H, Andersen NH, Gravholt CH. Causes of mortality in the marfan syndrome(from a nationwide register study). The American Journal of Cardiology. 2018;122:1231–1235

10. Lin XF, Xie LF, Zhang ZF, He J, Xie YL, Dai XF, et al. Quality of life in young patients with acute type a aortic dissection in china: Comparison with marfan syndrome and non-marfan syndrome. BMC Cardiovasc Disord. 2024;24:132

11. Dhanekula AS, Flodin R, Shibale P, Volk J, Benyakorn T, DeRoo S, et al. Natural history of the distal aorta following elective root replacement in patients with marfan syndrome. J Thorac Cardiovasc Surg. 2023

12. Quint LE, Liu PS, Booher AM, Watcharotone K, Myles JD. Proximal thoracic aortic diameter measurements at ct: Repeatability and reproducibility according to measurement method. Int J Cardiovasc Imaging. 2013;29:479–488

13. Lu TL, Rizzo E, Marques-Vidal PM, Segesser LK, Dehmeshki J, Qanadli SD. Variability of ascending aorta diameter measurements as assessed with electrocardiography-gated multidetector computerized tomography and computer assisted diagnosis software. Interact Cardiovasc Thorac Surg. 2010;10:217–221

14. Franken R, El Morabit A, de Waard V, Timmermans J, Scholte AJ, van den Berg MP, et al. Increased aortic tortuosity indicates a more severe aortic phenotype in adults with marfan syndrome. Int J Cardiol. 2015;194:7–12

15. Morris SA. Arterial tortuosity in genetic arteriopathies. Current opinion in cardiology. 2015;30:587–593

16. Sénémaud J, Gaudry M, Jouve E, Blanchard A, Milleron O, Dulac Y, et al. Primary non-aortic lesions are not rare in marfan syndrome and are associated with aortic dissection independently of age. J Clin Med. 2023;12

17. Ágg B, Szilveszter B, Daradics N, Benke K, Stengl R, Kolossváry M, et al. Increased visceral arterial tortuosity in marfan syndrome. Orphanet J Rare Dis. 2020;15:91

18. Morris SA, Orbach DB, Geva T, Singh MN, Gauvreau K, Lacro RV. Increased vertebral artery tortuosity index is associated with adverse outcomes in children and young adults with connective tissue disorders. Circulation. 2011;124:388–396

19. Spinardi L, Vornetti G, De Martino S, Golfieri R, Faccioli L, Pastore Trossello M, et al. Intracranial arterial tortuosity in marfan syndrome and loeys-dietz syndrome: Tortuosity index evaluation is useful in the differential diagnosis. AJNR Am J Neuroradiol. 2020;41:1916–1922

20. Yildiz M, Nucera M, Jungi S, Heinisch PP, Mosbahi S, Becker D, et al. Outcome of stanford type b dissection in patients with marfan syndrome. European Journal of Cardio-Thoracic Surgery. 2023;64

21. Lazea C, Bucerzan S, Crisan M, Al-Khzouz C, Miclea D, Şufană C, et al. Cardiovascular manifestations in marfan syndrome. Med Pharm Rep. 2021;94:S25–s27

22. Meester JAN, Verstraeten A, Schepers D, Alaerts M, Van Laer L, Loeys BL. Differences in manifestations of marfan syndrome, ehlers-danlos syndrome, and loeys-dietz syndrome. Ann Cardiothorac Surg. 2017;6:582–594

23. Bian Z, Zhong J, Dominic J, Christensen GE, Hatt CR, Burris NS. Validation of a robust method for quantification of three-dimensional growth of the thoracic aorta using deformable image registration. Med Phys. 2022;49:2514–2530

24. Burris NS, Lu Y, Knuer HA, Spahlinger G, Hatt CR. Abstract 11343: Unique phenotypes of three-dimensional aortic growth in genetic aortopathy. Circulation. 2022;146:A11343–A11343

25. Burris NS, Bian Z, Dominic J, Zhong J, Houben IB, van Bakel TMJ, et al. Vascular deformation mapping for ct surveillance of thoracic aortic aneurysm growth. Radiology. 2022;302:218–225

26. Zhang J, Zhang A, Wang Z, Sun Y, Li X, Jin Q, et al. A comparative study of clinical and aortic morphological characteristics between bovine aortic arch and normal aortic arch in patients with acute type b aortic dissection. Cardiology. 2023;148:409–417

27. Katakol S, Baker TJ, Bian Z, Lu Y, Spahlinger G, Hatt CR, et al. Fully automated pipeline for measurement of the thoracic aorta using joint segmentation and localization neural network. Journal of Medical Imaging. 2023;10

28. Liu X. Classification accuracy and cut point selection. Stat Med. 2012;31:2676–2686

29. Lopez-Sainz A, Mila L, Rodriguez-Palomares J, Limeres J, Granato C, La Mura L, et al. Aortic branch aneurysms and vascular risk in patients with marfan syndrome. J Am Coll Cardiol. 2021;77:3005–3012

30. Sun L, Li X, Wang G, Sun J, Zhang X, Chi H, et al. Relationship between length and curvature of ascending aorta and type a dissection. Front Cardiovasc Med. 2022;9:927105

31. Eliathamby D, Gutierrez M, Liu A, Ouzounian M, Forbes TL, Tan KT, et al. Ascending aortic length and its association with type a aortic dissection. J Am Heart Assoc. 2021;10:e020140

32. Zhang X, Peng Y, Li G, Li J, Luo M, Che Y, et al. Elongation of the proximal descending thoracic aorta and associated hemodynamics increase the risk of acute type b aortic dissection. Technol Health Care. 2023

33. Wu J, Zafar MA, Li Y, Saeyeldin A, Huang Y, Zhao R, et al. Ascending aortic length and risk of aortic adverse events: The neglected dimension. J Am Coll Cardiol. 2019;74:1883–1894

34. Gulati A, Zamirpour S, Leach J, Khan A, Wang Z, Xuan Y, et al. Ascending thoracic aortic aneurysm elongation occurs in parallel with dilatation in a nonsurgical population. Eur J Cardiothorac Surg. 2023;63

35. Morris SA, Orbach DB, Geva T, Singh MN, Gauvreau K, Lacro RV. Increased vertebral artery tortuosity index is associated with adverse outcomes in children and young adults with connective tissue disorders. Circulation. 2011;124:388–396

36. Narula N, Devereux RB, Arbustini E, Ma X, Weinsaft JW, Girardi L, et al. Risk of type b dissection in marfan syndrome: The cornell aortic aneurysm registry. J Am Coll Cardiol. 2023

37. Holmes KW, Maslen CL, Kindem M, Kroner BL, Song HK, Ravekes W, et al. Gentac registry report: Gender differences among individuals with genetically triggered thoracic aortic aneurysm and dissection. Am J Med Genet A. 2013;161a:779–786

38. Jones KB, Myers L, Judge DP, Kirby PA, Dietz HC, Sponseller PD. Toward an understanding of dural ectasia: A light microscopy study in a murine model of marfan syndrome. Spine (Phila Pa 1976). 2005;30:291–293

39. Engelfriet PM, Boersma E, Tijssen JG, Bouma BJ, Mulder BJ. Beyond the root: Dilatation of the distal aorta in marfan’s syndrome. Heart. 2006;92:1238–1243

40. Nannini G, Caimi A, Palumbo MC, Saitta S, Girardi LN, Gaudino M, et al. Aortic hemodynamics assessment prior and after valve sparing reconstruction: A patient-specific 4d flow-based fsi model. Comput Biol Med. 2021;135:104581

41. Guala A, Rodriguez-Palomares J, Dux-Santoy L, Teixido-Tura G, Maldonado G, Galian L, et al. Influence of aortic dilation on the regional aortic stiffness of bicuspid aortic valve assessed by 4-dimensional flow cardiac magnetic resonance. JACC: Cardiovascular Imaging. 2019;12:1020–1029

42. Park R, Latvis C, Roman M, Kim J, Agoglia H, Liberman N, et al. Aortic strain in patients with marfan syndrome long-term after proximal graft replacement surgery – novel insights enabled by cine-cmr biomechanical characterization. 2022

43. Petsophonsakul P, Furmanik M, Forsythe R, Dweck M, Schurink GW, Natour E, et al. Role of vascular smooth muscle cell phenotypic switching and calcification in aortic aneurysm formation. Arterioscler Thromb Vasc Biol. 2019;39:1351–1368

44. Kosciuszek ND, Kalta D, Singh M, Savinova OV. Vitamin k antagonists and cardiovascular calcification: A systematic review and meta-analysis. Front Cardiovasc Med. 2022;9:938567

45. Kälsch H, Lehmann N, Moebus S, Hoffmann B, Stang A, Jöckel KH, et al. Aortic calcification onset and progression: Association with the development of coronary atherosclerosis. Journal of the American Heart Association. 2017;6:e005093

46. Wanga S, Hibender S, Ridwan Y, van Roomen C, Vos M, van der Made I, et al. Aortic microcalcification is associated with elastin fragmentation in marfan syndrome. J Pathol. 2017;243:294–306

47. Yetman AT, McCrindle BW. The prevalence and clinical impact of obesity in adults with marfan syndrome. Can J Cardiol. 2010;26:137–139

